# Translation, adaptation, and validation of the Hospital Consumer Assessment of Healthcare Providers and Systems (HCAHPS) for use in Japan: A multicenter cross-sectional study

**DOI:** 10.1101/2020.05.14.20102442

**Authors:** Takuya Aoki, Yosuke Yamamoto, Tomoaki Nakata

## Abstract

**Objectives:** The Hospital Consumer Assessment of Healthcare Providers and Systems (HCAHPS) is a well-established and internationally recognized scale for measuring patients’ experience with hospital inpatient care. This study aimed to develop a Japanese version of the HCAHPS and to examine its structural validity, criterion-related validity, and internal consistency reliability.

**Design:** Multicenter cross-sectional study.

**Setting:** A total of 48 hospitals in Japan.

**Participants:** Patients aged ≥ 16 years who were discharged from the participating hospitals.

**Results:** We translated the HCAHPS into Japanese according to the guidelines. Psychometric properties were examined using data from 6,522 patients. A confirmatory factor analysis showed excellent goodness of fit of the same factor structure as that of the original HCAHPS, with the following composites: communication with nurses, communication with doctors, responsiveness of hospital staff, hospital environment, communication about medicines, and discharge information. All hospital-level Pearson correlation coefficients between the Japanese HCAHPS composites and overall hospital rating exceeded the criteria. Results of inter-item correlations indicated adequate internal consistency reliability.

**Conclusions:** We developed the Japanese HCAHPS, and evaluated its structural validity, criterion-related validity, and internal consistency reliability. This scale could be used for quality improvement based on the assessment of patients’ experience with hospital care and for health services research in Japan.

**Strengths and limitations of this study:**

- The Japanese HCAHPS is the first validated scale measuring patients’ experience with hospital inpatient care in Japan.
- Our data were collected from a large number of hospitals that were distributed widely throughout Japan, and covered various hospital sizes and regions.
- Although we examined the structural validity, criterion-related validity, and internal consistency reliability of the Japanese version developed in this study, other psychometric properties, including convergent and discriminant validity, test-retest reliability, and interpretability, have not been assessed.

## INTRODUCTION

In recent years, better patients’ perceptions of quality of health care have been deemed as one of the crucial goals of health care. Thus, patient experience has been globally considered as an important quality indicator in a wide range of settings.[1, 2] Patient experience is integrally tied to the principles and practices of patient- and family-centered care. Embedded within patient experience is a focus on individualized care, and tailoring services to meet patients’ needs and engage them as partners in their care.[3] Patient experience has recently replaced patient satisfaction because there are some limitations regarding the assessment of patient satisfaction, such as poor discriminability.[4] Several studies have shown that patient experience is consistently positively associated with clinical effectiveness, patient safety, and patient behaviors across a wide range of disease areas, settings, population groups, and outcome measures.[5–8].

The Hospital Consumer Assessment of Healthcare Providers and Systems (HCAHPS) is a well-established and internationally recognized scale for measuring patients’ experience with hospital care.[9] This scale was developed by the Centers for Medicare and Medicaid Services (CMS) in partnership with and funded by the Agency for Healthcare Research and Quality (AHRQ).[10] In the United States, HCAHPS results have been linked to financial reimbursement from Medicare and other insurers for promoting quality improvement in hospitals.[11] Additionally, these results are posted on the website for helping patients’ decision-making process by enabling comparisons across hospitals.[12]

In Japan, activities for assessment of patient experience have just begun in limited settings, and systematic approaches for quality improvement based on patients’ perceptions of health care are still inadequate. In recent years, several scales have been developed and validated to assess outpatients’ experience, mainly in the primary care setting.[13–15] However, there are no validated scales for assessing patients’ experience with hospital inpatient care in Japan. Accordingly, the present study aimed to develop a Japanese version of the HCAHPS, and to examine its structural validity, criterion-related validity, and internal consistency reliability.

## METHODS

### Design, setting, and participants

This multicenter cross-sectional study was conducted in 48 hospitals from September to December 2019, in cooperation with the Nihon Hospital Alliance (NHA), which is a group purchasing organization in Japan. Since 2014, the NHA has conducted an annual patient experience survey to evaluate and improve patient-centeredness in hospitals in Japan.[16] The participating hospitals voluntarily participated in the present study. Table 1 shows the characteristics of the participating hospitals. These hospitals were distributed widely throughout Japan, covering both urban and rural areas. Majority of the hospitals were large (≥ 400 beds), publicly owned, general hospitals, and they had an intensive care unit (ICU). A self-administered questionnaire was distributed to patients aged ≥ 16 years who were discharged from one of the participating hospitals during the survey period. Patients who were unable to respond to the questionnaire due to severe physical or mental disorders were excluded. We collected completed surveys by mail.

**Table 1.** Hospital characteristics of 48 participating hospitals.

| Characteristic | Number (%) |
| --- | --- |
| Hospital size |  |
| Small (< 200 beds) | 4 (8.3) |
| Medium (200–399 beds) | 17 (35.4) |
| Large (≥ 400 beds) | 27 (56.3) |
| Ownership |  |
| Public | 34 (70.8) |
| Private | 14 (29.2) |
| Hospital type |  |
| General hospital | 46 (95.8) |
| Special hospital | 2 (4.2) |
| ICU |  |
| Yes | 29 (60.4) |
| No | 19 (39.6) |
| Hospital region |  |
| North | 10 (20.8) |
| East | 20 (41.7) |
| West | 18 (37.5) |
| Municipality population size |  |
| Small (< 50,000) | 6 (12.5) |
| Medium (50,000–200,000) | 14 (29.2) |
| Large (> 200,000) | 28 (58.3) |

### Measures

#### The HCAHPS

The original HCAHPS is a 19-item tool comprising six composites, two global ratings, and three screening items.[9] The composites are communication with nurses (Q1–Q3), communication with doctors (Q5–Q7), responsiveness of hospital staff (Q4 and 11), hospital environment (Q8 and 9), communication about medicines (Q13 and 14), and discharge information (Q16 and 17). The global ratings include overall hospital rating (Q18) and willingness to recommend the hospital to friends and family (recommended hospital) (Q19).

We obtained permission for translating the HCAHPS into Japanese from the AHRQ and CMS. According to the guidelines for translating CAHPS® surveys provided by the AHRQ,[17] we translated the HCAHPS into Japanese through the following steps. First, two forward translations from English to Japanese were performed independently by two bilingual translators who had prior professional experience in translating survey instruments for health services. Subsequently, the two forward translations were reviewed by a translation reviewer, who was a native speaker of Japanese and had prior experience in translating survey instruments. After reviewing the translations, the reviewer produced a reconciled version of the translation. The final version of the translation was then produced through discussion in a committee composed of the two translators and the reviewer. The reconciled version from the original review was modified as needed, based on the committee’s decision for cross-cultural adaptation. The final wording of each survey item and response option was determined by consensus.

The HCAHPS survey uses several different response scales: a dichotomous scale (1 = Yes, 2 = No), a global rating scale (0 = Worst to 10 = Best), and four-point Likert scales (1 = Never, 2 = Sometimes, 3 = Usually, and 4 = Always; and 1 = Definitely no, 2 = Probably no, 3 = Probably yes, and 4 = Definitely yes). To make the results easier to understand, we converted all scales to normalized scores ranging from 0 to 100 using the following formula:

Normalized Score = 100 * (Respondent’s selected response value – Minimum response value on the scale) / (Maximum response value – Minimum response value)

In the Japanese version, assuming the convergence in each composite as in the original version, the score for each of the six composites was computed as the mean value for all normalized scores in the scale that would fall in the range of 0–100 points, with higher scores indicating better performance.

### Statistical analysis

To validate the Japanese HCAHPS, we first conducted a confirmatory factor analysis to evaluate the structural validity of the Japanese HCAHPS composites. In the factor analysis, we hypothesized the same factor structure (six-factor solution) as that of the original HCAHPS. The appropriateness of the resulting structure was determined by examining if factor loadings were 0.40 or greater.[18] The model fitness was assessed by the comparative fit index (CFI), Tucker-Lewis index (TLI), root mean square error of approximation (RMSEA), and standardized root mean square residual (SRMR). Guidelines suggest that models with CFI and TLI close to 0.95 or higher, RMSEA close to 0.06 or lower, and SRMR close to 0.08 or lower are representative of models with a good fit.[19]

Subsequently, the Japanese HCAHPS composite scores and the overall hospital rating were used to examine criterion-related validity. Validity was assessed using Pearson correlation coefficients with each Japanese HCAHPS composite, to predict the overall hospital rating at the hospital level. A correlation coefficient greater than 0.30 was considered meaningful.[20] Hospital-level correlations are a more important criterion for measurement than are individual-level correlations because the former are benchmarking tools to compare one hospital with another. To examine hospital-level correlations, we used each hospital’s mean score on HCAHPS composites and the overall hospital rating.

The internal consistency reliability was examined by inter-item correlations and the Cronbach’s alpha. For a scale to be considered sufficiently reliable, an inter-item correlation of 0.30 and a Cronbach’s alpha value of 0.70 is recommended.[21] Finally, descriptive statistics were performed for the Japanese HCAHPS scores, including the mean, standard deviation, and observed range. To deal with missing data, in the confirmatory factor analysis, we used full information maximum likelihood estimation to enable use of information collected from participants with missing data. In the evaluation of criterion-related validity and internal consistency, we conducted complete case analyses. All statistical analyses were conducted using R version 3.6.3 (R Foundation for Statistical Computing, Vienna, Austria; www.R-project.org).

### Ethics approval

This study was approved by the Ethics Committee of Kyoto University Graduate School of Medicine (approval number R2331) and was conducted in accordance with the Declaration of Helsinki. We obtained consent for participation from each participant.

## RESULTS

Of the total 15,512 eligible participants, 6,522 (42.0%) responded to the survey. Table 2 shows the participants’ responses to each item of the Japanese HCAHPS. The Top Box score for each item, which is the percentage of participants who provided most positive responses on that item, ranged from 31.1% to 82.5%. Regarding the mean Top Box score for composites, the highest score was observed for discharge information (77.2%) while the lowest score was for hospital environment (49.1%). The bottom box score, which is the percentage of participants with least positive responses on the item, ranged from 0.5% to 23.2%.

**Table 2.**
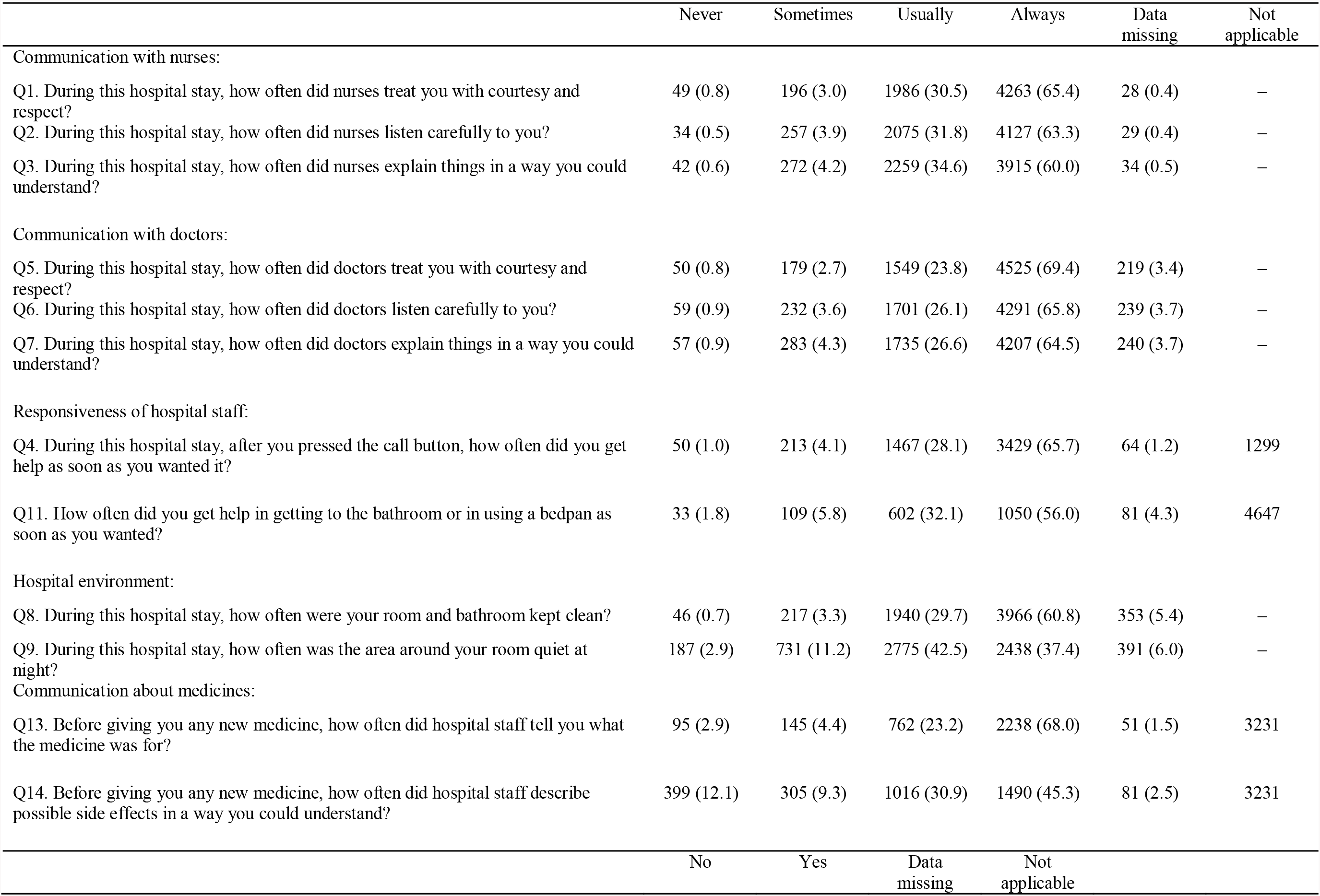
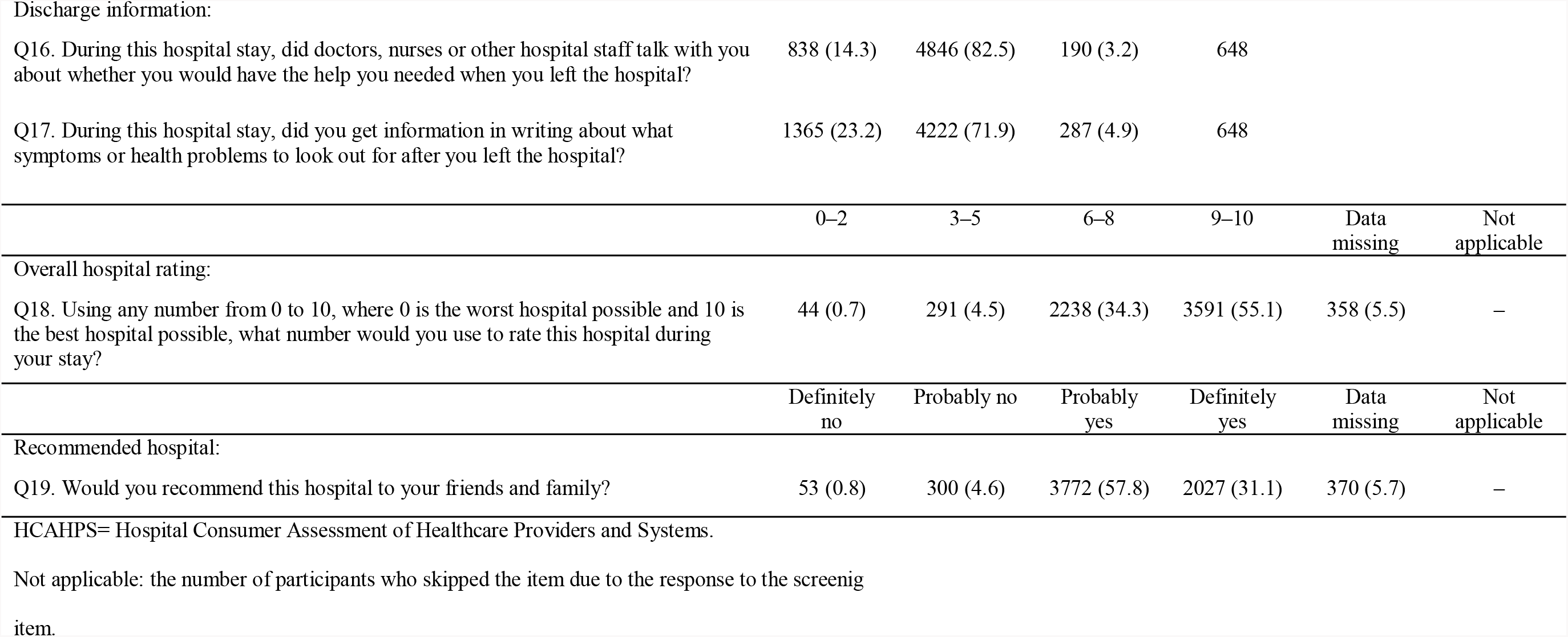
Response to Japanese HCAHPS items: number (%) (N = 6,522)

### Structural validity

Figure 1 shows the path diagrams of the confirmatory factor analysis to assess the structural validity of the Japanese HCAHPS composites. All factor loadings of each items onto each factor were above the 0.40 criteria, ranging from 0.41 to 0.91. The correlation coefficients among factors ranged from 0.30 to 0.85. The conceptual model showed excellent goodness of fit, with CFI = 0.987, TLI = 0.981, RMSEA = 0.031, and SRMR = 0.020.

**Figure 1.**
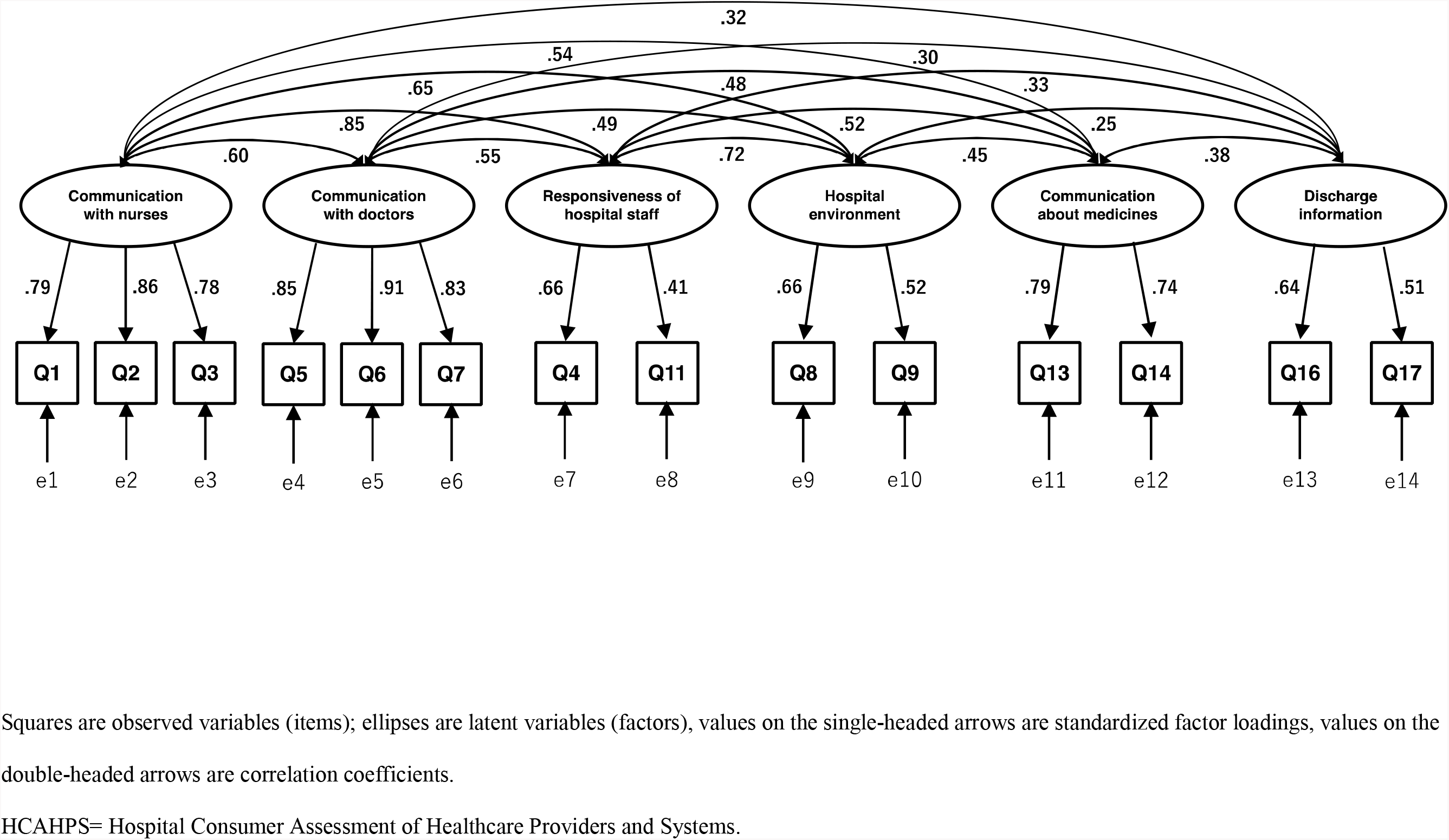
Factor structure of Japanese HCAHPS (confirmatory factor analysis)

### Criterion-related validity

Table 3 shows the Pearson correlation coefficients between the Japanese HCAHPS composites and the overall hospital rating at the hospital level. All correlations were statistically significant (*P* < 0.01), and they exceeded the 0.30 criterion. The composite “communication with doctors” (*r* = 0.63) had the highest correlation with the overall rating.

**Table 3.**
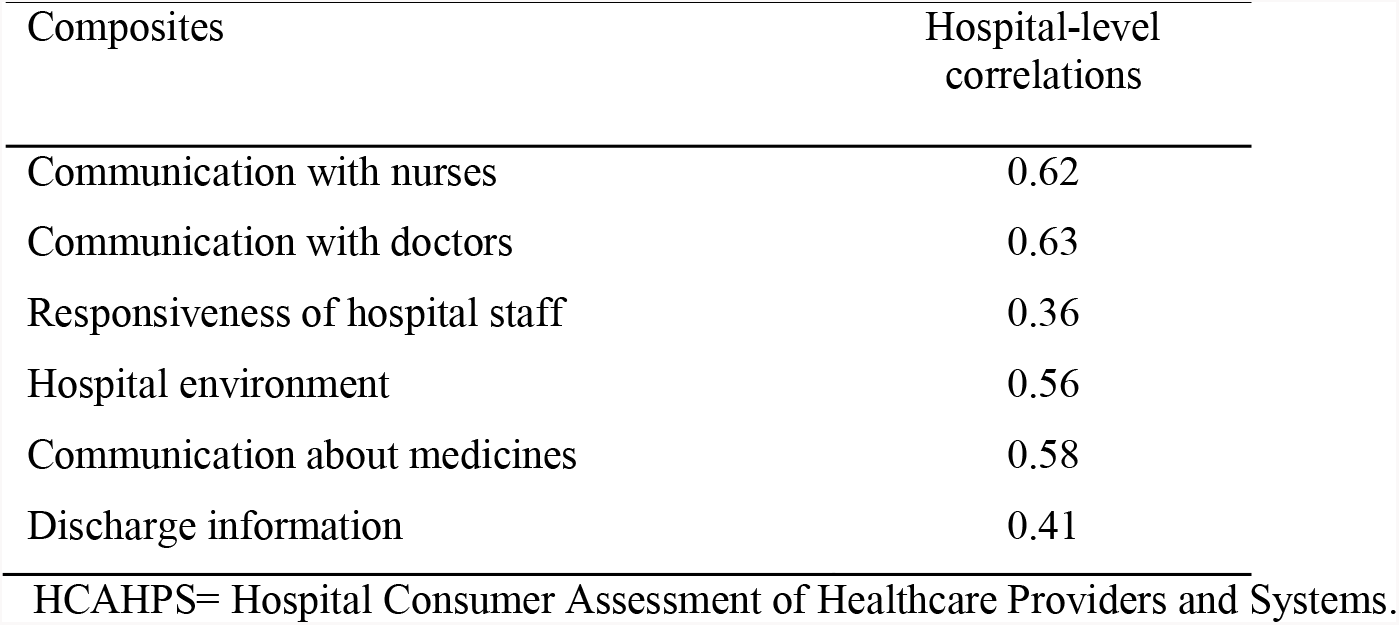
Pearson correlation coefficients between Japanese HCAHPS composites and overall hospital rating.

### Internal consistency reliability and descriptive statistics

Table 4 indicates the score distribution and internal consistency reliability for the Japanese HCAHPS. All inter-item correlations were above the 0.30 criteria, ranging from 0.31 to 0.73. For communication with nurses, communication with doctors, and communication about medicines, the Cronbach’s alpha was above 0.70. In contrast, for responsiveness of hospital staff, hospital environment, and discharge information, the Cronbach’s alpha did not exceed the 0.70 criterion. Descriptive statistics showed that the most highly-scored scale was communication with doctors (mean score = 87.9), and the most poorly-scored scale was recommended hospital (mean score = 75.5). The full range of possible scores was observed for all scales.

**Table 4.** Descriptive features and internal consistency reliability of Japanese HCAHPS (N = 6,522)

|  | Number of items | Mean | Standard deviation | Observed range | Inter-item correlation | Cronbach's alpha |
| --- | --- | --- | --- | --- | --- | --- |
| Composites |  |  |  |  |  |  |
| Communication with nurses | 3 | 86.1 | 17.4 | 0.0–100.0 | 0.62–0.73 | 0.85 |
| Communication with doctors | 3 | 87.9 | 18.1 | 0.0–100.0 | 0.65–0.70 | 0.89 |
| Responsiveness of hospital staff | 2 | 81.0 | 21.1 | 0.0–100.0 | 0.34 | 0.46 |
| Hospital environment | 2 | 80.2 | 18.7 | 0.0–100.0 | 0.35 | 0.49 |
| Communication about medicines | 2 | 78.7 | 25.8 | 0.0–100.0 | 0.61 | 0.71 |
| Discharge information | 2 | 80.3 | 32.0 | 0.0–100.0 | 0.31 | 0.48 |
| Global ratings |  |  |  |  |  |  |
| Overall hospital rating | 1 | 85.7 | 15.4 | 0.0–100.0 | – | – |
| Recommended hospital | 1 | 75.5 | 19.5 | 0.0–100.0 | – | – |

HCAHPS= Hospital Consumer Assessment of Healthcare Providers and Systems.

## DISCUSSION

Measurement of patient experience plays an important role in the improvement of a wide range of medical services, including inpatient care. We translated the HCAHPS, which is a validated international scale, into Japanese and examined its structural validity, criterion-related validity, and internal consistency reliability in 48 hospitals in Japan. This study was the first to develop a Japanese version of the HCAHPS and to examine its psychometric properties for assessing patients’ experience with hospital inpatient care.

Standard psychometric evaluation methods were used to evaluate the Japanese HCAHPS. The confirmatory factor analysis supported the scale’s structural validity and the same six-factor solution as that of the original HCAHPS, with good statistical fitness. Correlation coefficients between all Japanese HCAHPS composites and the overall hospital rating for assessing criterion-related validity exceeded the meaningful value at the hospital level.

In internal consistency analyses, the Cronbach’s alpha for responsiveness of hospital staff, hospital environment, and discharge information did not exceed the optimum criterion. The Cronbach’s alpha is quite sensitive to the number of items in the scale; therefore, it is common to find low Cronbach’s alpha for scales with few items (especially 2-item scales).[22] Likewise, a study conducted in the United States [23] found that the Cronbach’s alpha was low for some aspects of the original HCAHPS scale. In this case, it is more appropriate to report the inter-item correlation of items. In our study, all inter-item correlations were greater than the criterion, which indicated adequate internal consistency of the scales.

To our knowledge, the Japanese HCAHPS is the first validated scale measuring patients’ experience with hospital inpatient care in Japan. The HCAHPS is one of the most widely studied and endorsed patient experience measure of hospital care worldwide. Our data were collected from a large number of hospitals that were distributed widely throughout Japan and they covered various hospital sizes and regions. Therefore, the study results have relatively high external validity.

However, there are several potential limitations to our study. First, the response rate was a concern. A previous study of patient experience surveys showed that a low participation rate is less likely to introduce selective nonresponse bias;[24] however, it is possible that patients with worse experience were less likely to respond to our survey. Second, although we examined the structural validity, criterion-related validity, and internal consistency reliability of the Japanese HCAHPS in this study, other psychometric properties, including convergent and discriminant validity, test-retest reliability, and interpretability have not been assessed.[25] These measurement properties of the scale need to be evaluated in future studies. Third, this study was limited by the fact that the participating hospitals voluntarily participated in this study; thus, the preset sample may represent hospitals that have a higher interest in the quality of health care. Accordingly, the participating hospitals may not have sufficiently represented Japanese hospitals at national level. Therefore, the Japanese HCAHPS should be used for research in other settings.

## CONCLUSION

We developed the Japanese HCAHPS and evaluated its structural validity, criterion-related validity, and internal consistency reliability. This scale could be used for quality improvement based on the assessment of patients’ experience with hospital care and for health services research in Japan.

## Data Availability

Additional unpublished data is still being analyzed for another research and only available to the members of the study team.

## Acknowledgments

We would like to thank Toshio Goto and Yoko Endo (Nihon Hospital Alliance) for their assistance with the administration of this study. We are grateful to Prof. Kiyoshi Ando, Prof. Keiichi Saito, and Kaori Soga (Association for Patient eXperience Japan) for collaboration on the early stages of this work.

## Contributors

TA designed the study and participated in the implementation, data analysis and writing of the manuscript. YY contributed to the design of the study and critically reviewed the manuscript. TN contributed to the design of the study, data collection, and critically reviewed the manuscript. All authors gave the final approval of the manuscript before submission.

## Patient consent for publication

Not required.

## Patient and Public Involvement statement

We did not directly include PPI in this study, but the database used in the study was developed with PPI.

